# Influenza vaccine effectiveness against hospitalized SARS-CoV-2 infection

**DOI:** 10.1101/2023.10.27.23297682

**Authors:** Yung-Chun Lee, Lufeiya Liu, Liyang Yuan, Malcolm Risk, Kevin Heinrich, Martin Witteveen-Lane, Salim Hayek, Ryan Malosh, Rodica Pop-Busui, Bian Jiang, Chen Shen, Dave Chesla, Richard Kennedy, Shi Xu, Matthew Sims, Ramin Homayouni, Lili Zhao

**Affiliations:** Division of Biostatistics & Health Informatics, Beaumont Research Institute, Corewell Health East, Royal Oak, MI 48073; Department of Biostatistics, University of Michigan, Ann Arbor, MI 48109; Quire Data, Royal Oak, MI 48073; Bioinformatics Core, Corewell Health Wast, Grand Rapids, MI 49503; Division of Cardiology, Department of Medicine, University of Michigan, Ann Arbor, MI 48109; Division of Immunizations, MDHHS, Lansing, MI 48909; Department of Internal Medicine, University of Michigan, Ann Arbor, MI 48105; College of Medicine Division of Health Outcome & Biomedical Informatics, University of Florida Health, Gainesville, FL 32608; Beaumont Research Institute, Corewell Health East, Royal Oak, MI 48073; Department of Infectious Disease, Corewell Health East, Royal Oak, MI 48073; Department of Foundational Medical Studies, OUWB School of Medicine, Rochester, MI 48309

**Keywords:** Influenza vaccine, COVID-19 vaccine, Covid-19 Infection, Covid-19 hospitalization

## Abstract

**Background:** Some studies conducted before the Delta and Omicron variant-dominant periods have indicated that influenza vaccination provided protection against COVID-19 infection or hospitalization, but these results were limited by small study cohorts and a lack of comprehensive data on patient characteristics. No studies have examined this question during the Delta and Omicron periods (08/01/2021 to 2/22/2022).

**Methods:** We conducted a retrospective cohort study of influenza-vaccinated and unvaccinated patients in the Corewell Health East(CHE, formerly known as Beaumont Health), Corewell Health West(CHW, formerly known as Spectrum Health) and Michigan Medicine (MM) healthcare system during the Delta-dominant and Omicron-dominant periods. We used a test-negative, case-control analysis to assess the effectiveness of the influenza vaccine against hospitalized SARS-CoV-2 outcome in adults, while controlling for individual characteristics as well as pandameic severity and waning immunity of COVID-19 vaccine.

**Results:** The influenza vaccination has shown to provided some protection against SARS-CoV-2 hospitalized outcome across three main healthcare systems. CHE site (odds ratio [OR]=0.73, vaccine effectiveness [VE]=27%, 95% confidence interval [CI]: [18-35], p<0.001), CHW site (OR=0.85, VE=15%, 95% CI: [6-24], p<0.001), MM (OR=0.50, VE=50%, 95% CI: [40-58], p <0.001) and overall (OR=0.75, VE=25%, 95% CI: [20-30], p <0.001).

**Conclusion:** The influenza vaccine provides a small degree of protection against SARS-CoV-2 infection across our study sites.

## 1 Introduction

Due to the constantly changing nature of influenza viruses, flu shots are given on a seasonal basis, with around 53% of adults in Michigan opting to receive the flu shot each year[1]. The flu shot is typically between 30% and 50% effective against influenza infection, depending on the study population and season[2]. There has been some speculation that the flu shot may also provide some protection against SARS-CoV-2 infection and COVID-19 disease[3]. Researchers have proposed several mechanisms, such as trained immunity[4][5][6][7][8][9][10][11], increased cytokine production[12][13], and viral interference[14], to explain how heterologous or nonspecific effects (NSEs) induced by available non–SARS-CoV-2 vaccines could potentially prevent SARS-CoV-2 infection and reduce COVID-19 disease.[15]

Despite some small studies have shown that there is not enough evidence to claim that influenza vaccination has no efficacy against Covid-19 during the beginning of the global pandemic before delta and omicron-variant[16][17][18], more studies conducted with a larger sample size suggested differently including studies conducted in Brazil[19], Turkey[20], Canada[21], Italy[22][23], and other states within the US[24][25][26]. A research group from Italy [22] has found that anti-pneumococcal and influenza vaccinations are associated with a lower positive test rate in younger group and even lower rate in elderly group using web-based survey data. However, there is still a large degree of uncertainty in the level of protection provided by the flu vaccine. Studies with a limited ability to control for patient characteristics are likely to overstate influenza vaccine effectiveness because people who received vaccination have better health-seeking behavior in general. They might take health-seeking choices, such as masking, social distancing, COVID-19 vaccination, or have routine health visits and follow doctor’s order and recommendation. Several Studies used survey data [22][24][27], but it might subject to selection bias and respondents are not representative of the real-world population regardless of the large sample size. In addition, many studies were conducted using data collected at the beginning of global pandemic in 2020 before other highly transmissible variants, such as delta variant, were introduced, and more studies using more recent data are needed.

In this study, we aimed to estimate influenza vaccine (flu shot) effectiveness against the hospitalized SARS-CoV-2 over the spam of Delta and Omicron periods, using electronic health record (EHR) from three largest healthcare system in the state of Michigan (CHE, CHW and MM).

## 2 Methods

### 2.1 Data

We included patients who were registered with a primary care physician were included and had at least one primary care physician (PCP) visit within 18 months prior to 2021/1/1 and had at least one SARS-CoV-2 molecular (PCR) test done within study period (08/01/2021 ∼02/22/2022).

We further excluded individuals who are under 18 on 2021/1/1, received a different COVID-19 vaccine than *Pfizer* or *Moderna*, or have incomplete covariate data. We used the same inclusion and exclusion criteria across all 3 sites, MM, CHE and CHW. The final number of subjects included in the analysis for the covid variant study period are 37,201 at MM, 70,486 at CHE and 102,467 at CHW site (see Table 1).

**Table 1:** Data Source across sites and number of eligible patients.

|  | MM | CHE | CHW |
| --- | --- | --- | --- |
| EHR(Epic) | ✓ | ✓ | ✓ |
| MCIR(Influenza) | ✓ | ✓ |  |
| MCIR(Covid-19) | ✓ | ✓<sup>1</sup> |  |
| Final number | 37,201 | 70,486 | 102,467 |

Michigan Care Improvement Registry (MCIR) documents immunizations given to Michigan residents throughout life and includes all immunizations statewide. All of our study site databases are linked with MCIR system, patients’ immunization records will be transferred and integrated into respective healthcare providers EHR when patient visit the hospital or visit. Comorbidities were assessed based on diagnosis codes 12 months before our study period on 08/01/2021.

We identified hospitalized SARS-CoV-2 cases for MM based on chart reviewed data, for CHE and CHW sites we used a similar criteria from a previous bivalent covid-19 vaccine efficacy study [**?**] for defining hospitalized COVID-19 outcome using hospitalization in conjunction with COVID-19 PCR test results, of which Emergency Department encounters and hospitalization associated with COVID-19 illness and have positive PCR test during 14 days hospitalization stay and 3 days prior to admission are considered as hospitalized COVID-19 cases.

Our primary interest is to study influenza vaccine effect against hospitalized COVID-19 outcome over the spam of Delta and Omicron variants periods against hospitalized COVID-19 outcome. We have determined the disease status based on PCR test results during the study period. If an individual had a positive PCR test, then this subject had the hospitalized infection and the date of the first positive PCR was used as the date of hospitalized infection. If an individual only had negative PCR test result(s), this subject did not have hospitalized infection and the date of the last test was used as the date of non-infection, see Figure 1. In this way, we each individual is classified into one disease status(hospitalized COVID-19 or no hospitalized COVID). We considered subjects to be vaccinated for influenza if they received an influenza vaccine within one year of PCR test date, and not vaccinated otherwise. We only evaluated the influenza vaccines’ effectiveness against hospitalized COVID-19 case during 08/01/2021 ∼02/22/2022.

**Figure 1:**
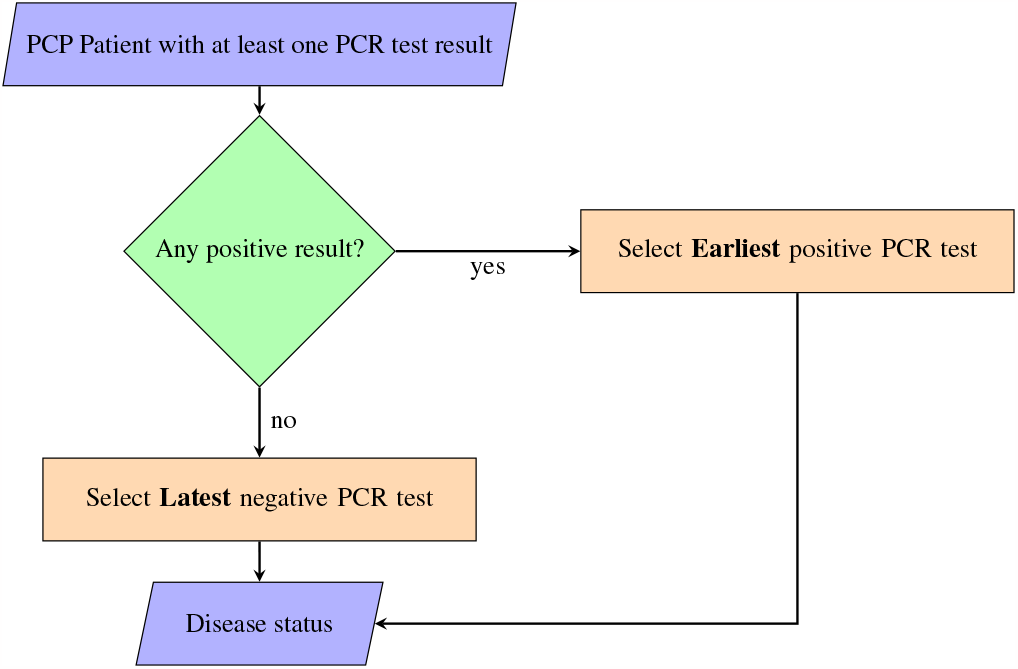
Disease Status classification

### 2.2 Statistical Analysis

We first conducted separate analysis for each study site using the same covariates, study design and statistical model. All analysis was conducted using complete cases after removing patients with missing covariates.

Logistic regression was used to study the effect of influenza vaccination against hospitalized SARS-CoV-2 infection by comparing the odds of prior influenza vaccination (exposed) vs. no vaccination (unexposed) in cases (hospitalized COVID-19 outcome cases) vs. controls (other PCR takers). Logistic regression models were adjusted for age, race, gender, and Charlson comorbidity index (CCI), calendar time of the PCR test (PCR Time: a categorical variable), and time from the last COVID-19 vaccine (a categorical variable, requiring at least 14 days from the most recent dose). PCT Test was used to control for pandemic severity over the study period. Due to waning immunity over time, we have previously found that number of doses is not associated with COVID-19 infection after controlling for time from last COVID-19 vaccine. We therefore categorized time from last COVID-19 vaccine into 5 categories: ‘Unvaccinated’, ‘less than 90 days’, ‘90 ∼180 days’, ‘180 days to one year’, and ‘more than one year’ to adjust and examine the waning effect of COVID-19 immunization. 28 Calendar week dates was divided into two-week periods to account for varying infection rate of COVID-19 which included but not limited to high exposure rate due to national holidays. The CCI was obtained using the R package comorbidity and categorized into 4 categories, ‘0’, ‘1-2’, ‘3-4’, and ‘≥ 5’. [28][29]. We reported Odds Ratio (OR), 95% Confidence Interval (CI) and p-value. The vaccine effectiveness (VE) was defined as VE% = (1-OR)%×100%

We used the Renewable estimation method [30] to combine EHR data from CHE, CHW and MM in a privacy-preserving way. The method is built upon sequentially updating regression coefficient estimates at each site such that the final estimate corresponds to what we would have found after pooling all our data. This method has been shown to achieve performance very close to that combines individual patient-level data from three sites. In our study, we started with MM and generated summary statistics (i.e., regression coefficients and corresponding Hessian matrix) and passes them to CHE, these summary statistics received from MM serve as “prior” for regression parameters and are combined with CHE data to form an updated “posterior” estimates for the regression coefficients. We then repeated this procedure to combine with CHW data to obtain the final estimates.

Statistical analyses were performed in R 4.2.3. OR and VE estimates were presented with 95% CIs and p-value. We used de-identified EHR data, the use of which was approved by the Institutional Review Board (IRB) of each sites.

## 3 Results

### 3.1 Study Population

There were 18,468 (26.2%) flu-vaccinated subjects from CHE site, 23,306 (22.7%) from CHW site, and 25,272 (67.9%) from MM during the study period. We observed that MM site has a higher rate in both influenza and COVID-19 immunization than other 2 sites.

Subjects at CHE site are older than the other CHW and MM sites. Median age of 56 (interquartile range [IQR]: 38-70) in CHE compared to 47 (IQR: 32-64) in CHW and 49 (IQR: 33-65) in MM.

Gender distribution are similar across 3 sites. CHE has 63.1% female, and 68.6% white, CHW site has 64.1% female, and 85.9% white and MM site has 61.2% female, and 76.9% white.

Overall, individuals who took the influenza vaccines were more likely to take the COVID-19 vaccine (see Figures 2). In our study period, among 210,136 subjects, 31.9% were flu-vaccinated before the PCR test, of whom 14.2% (had not received the COVID-19 vaccine. 68.1% of subjects were not flu-vaccinated, of whom 46.9% had not received the COVID-19 vaccine(see Figure 2). Patient covariates by hospitalized covid outcome are described in Table 2.

**Table 2:**
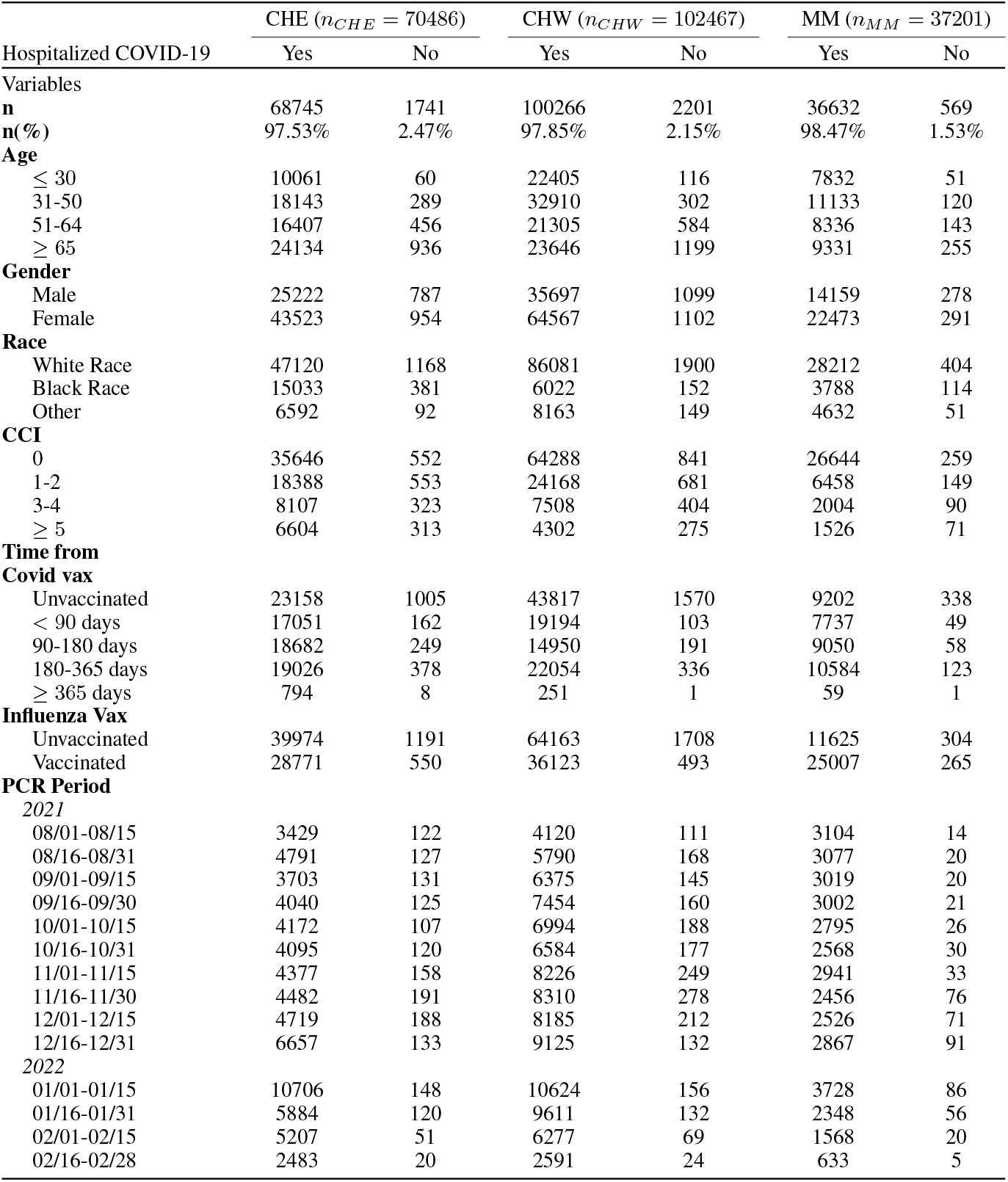
Study Population Descriptive Table.

| | CHE ($n_{CHE} = 70486$) | | CHW ($n_{CHW} = 102467$) | | MM ($n_{MM} = 37201$) | |
| --- | --- | --- | --- | --- | --- | --- |
| Hospitalized COVID-19 | Yes | No | Yes | No | Yes | No |
| **Variables** |  |  |  |  |  |  |
| **n** | 68745 | 1741 | 100266 | 2201 | 36632 | 569 |
| **n(%)** | 97.53% | 2.47% | 97.85% | 2.15% | 98.47% | 1.53% |
| **Age** |  |  |  |  |  |  |
| ≤ 30 | 10061 | 60 | 22405 | 116 | 7832 | 51 |
| 31-50 | 18143 | 289 | 32910 | 302 | 11133 | 120 |
| 51-64 | 16407 | 456 | 21305 | 584 | 8336 | 143 |
| ≥ 65 | 24134 | 936 | 23646 | 1199 | 9331 | 255 |
| **Gender** |  |  |  |  |  |  |
| Male | 25222 | 787 | 35697 | 1099 | 14159 | 278 |
| Female | 43523 | 954 | 64567 | 1102 | 22473 | 291 |
| **Race** |  |  |  |  |  |  |
| White Race | 47120 | 1168 | 86081 | 1900 | 28212 | 404 |
| Black Race | 15033 | 381 | 6022 | 152 | 3788 | 114 |
| Other | 6592 | 92 | 8163 | 149 | 4632 | 51 |
| **CCI** |  |  |  |  |  |  |
| 0 | 35646 | 552 | 64288 | 841 | 26644 | 259 |
| 1-2 | 18388 | 553 | 24168 | 681 | 6458 | 149 |
| 3-4 | 8107 | 323 | 7508 | 404 | 2004 | 90 |
| ≥ 5 | 6604 | 313 | 4302 | 275 | 1526 | 71 |
| **Time from Covid vax** |  |  |  |  |  |  |
| Unvaccinated | 23158 | 1005 | 43817 | 1570 | 9202 | 338 |
| < 90 days | 17051 | 162 | 19194 | 103 | 7737 | 49 |
| 90-180 days | 18682 | 249 | 14950 | 191 | 9050 | 58 |
| 180-365 days | 19026 | 378 | 22054 | 336 | 10584 | 123 |
| ≥ 365 days | 794 | 8 | 251 | 1 | 59 | 1 |
| **Influenza Vax** |  |  |  |  |  |  |
| Unvaccinated | 39974 | 1191 | 64163 | 1708 | 11625 | 304 |
| Vaccinated | 28771 | 550 | 36123 | 493 | 25007 | 265 |
| **PCR Period** |  |  |  |  |  |  |
| **2021** |  |  |  |  |  |  |
| 08/01-08/15 | 3429 | 122 | 4120 | 111 | 3104 | 14 |
| 08/16-08/31 | 4791 | 127 | 5790 | 168 | 3077 | 20 |
| 09/01-09/15 | 3703 | 131 | 6375 | 145 | 3019 | 20 |
| 09/16-09/30 | 4040 | 125 | 7454 | 160 | 3002 | 21 |
| 10/01-10/15 | 4172 | 107 | 6994 | 188 | 2795 | 26 |
| 10/16-10/31 | 4095 | 120 | 6584 | 177 | 2568 | 30 |
| 11/01-11/15 | 4377 | 158 | 8226 | 249 | 2941 | 33 |
| 11/16-11/30 | 4482 | 191 | 8310 | 278 | 2456 | 76 |
| 12/01-12/15 | 4719 | 188 | 8185 | 212 | 2526 | 71 |
| 12/16-12/31 | 6657 | 133 | 9125 | 132 | 2867 | 91 |
| **2022** |  |  |  |  |  |  |
| 01/01-01/15 | 10706 | 148 | 10624 | 156 | 3728 | 86 |
| 01/16-01/31 | 5884 | 120 | 9611 | 132 | 2348 | 56 |
| 02/01-02/15 | 5207 | 51 | 6277 | 69 | 1568 | 20 |
| 02/16-02/28 | 2483 | 20 | 2591 | 24 | 633 | 5 |

**Figure 2:**
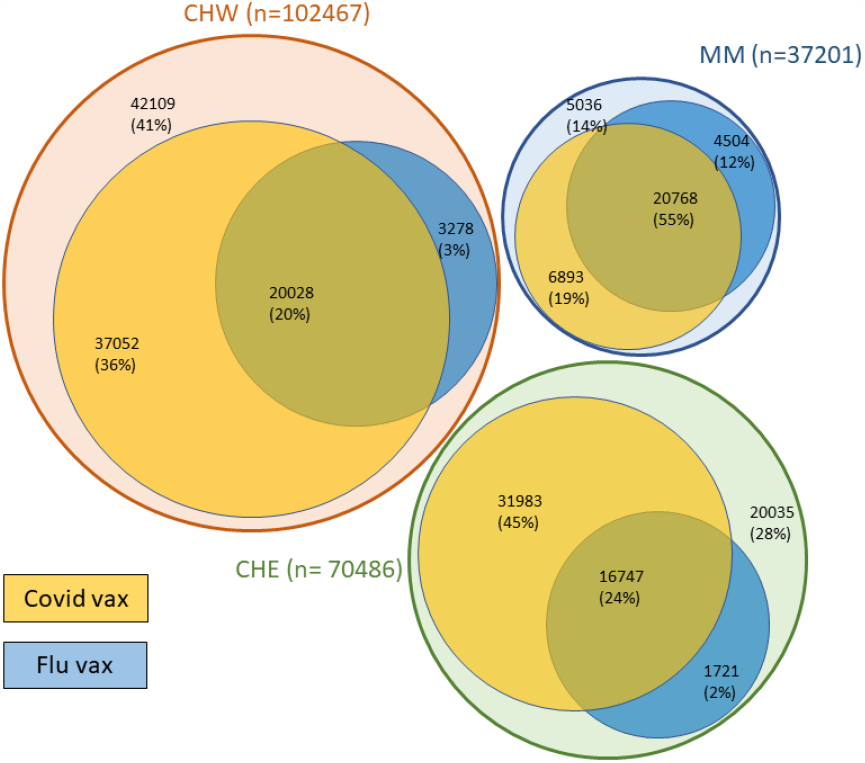
Influenza Vaccine and Covid Vaccine Venn Diagram

### 3.2 Influenza Vaccine Effectiveness against hospitalized SARS-CoV-2 Infection

We found that taking a influenza vaccine provided moderate protection against hospitalized SARS-CoV-2 infection (see Figure 3) in over Delta and Omicron-variant periods across all sites. For CHE site, influenza vaccines has shown to have lower OR against hospitalized COVID-19 outcome compared to those were not vaccinated within influenza vaccine(OR=0.73, CI:[0.65-0.82], VE=23%, p-value <0.001). Same results were found for CHW site(OR=0.85, CI:[0.76-0.94], VE=15%, p-value <0.001) and MM site(OR=0.50, CI:[0.42-0.60], VE=50%, p-value <0.001). The overall OR using the renewable estimation method from all study sites remained significant (OR=0.75, CI:[0.70-0.80], VE=25%, p<0.001).

**Figure 3:**
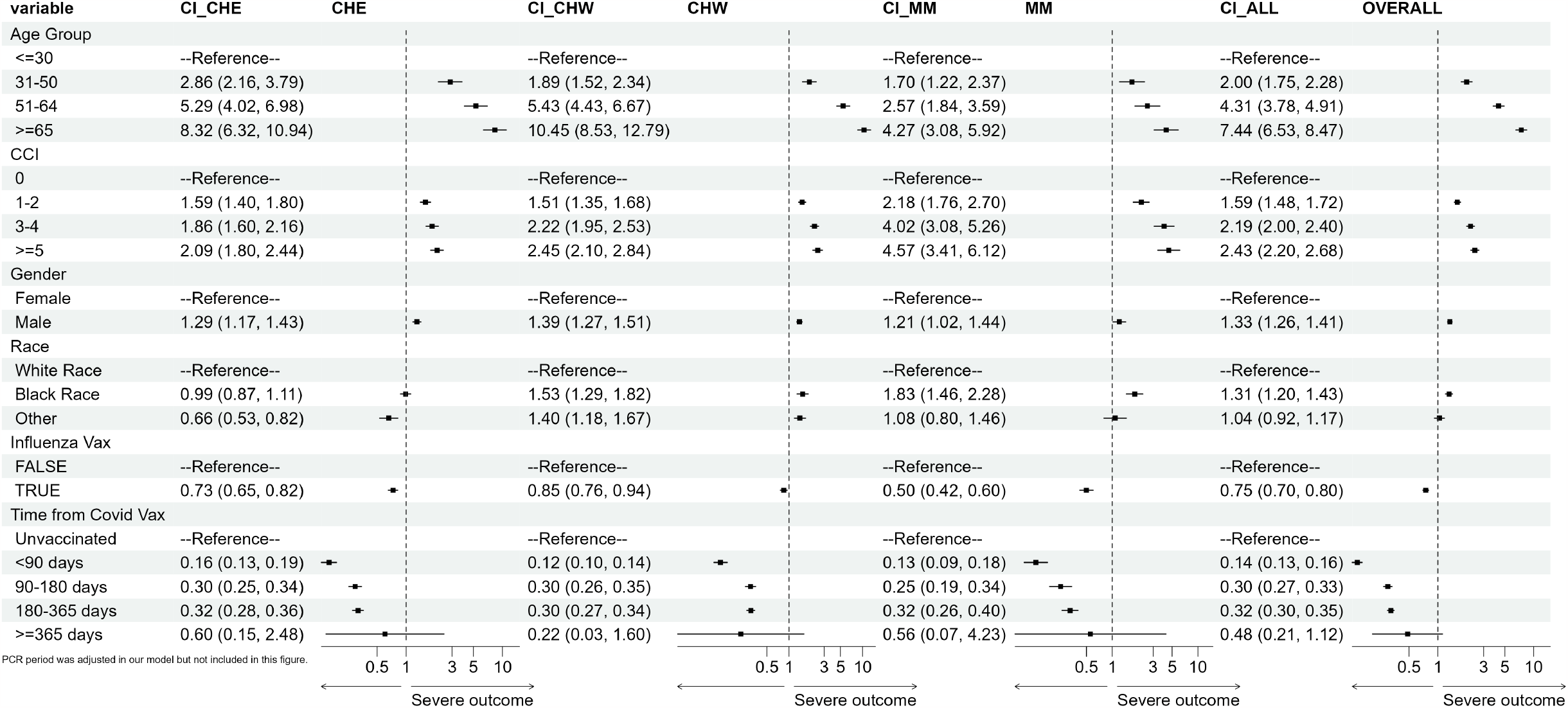
Odds Ratio by Site and integrated inference

### 3.3 COVID-19 Vaccine Effectiveness against hospitalized SARS-CoV-2 Infection

Receiving the COVID-19 vaccine more recently provided some protection against COVID-19 infection in Covid variant period, and we also observed the waning effect of COVID-19 vaccine compared to those who are not vaccinated with COVID-19 vaccine.

For CHE site, the adjusted OR for individuals having received their most recent COVID-19 vaccine within 90 days was 0.16 (VE =84%, 95% CI: [81-87]), for individuals having received their most recent COVID-19 vaccine 90-180 days ago was 0.30 (VE =70%, 95% CI: [66-75]), for individuals having received the most recent COVID-19 vaccine 180-365 days was 0.32 (VE =68%, 95% CI: [64-72]), subjects who took their last COVID-19 vaccine more than 365 days earlier was not significant (OR= 0.60, p=0.48)in our study period.

Same waning patterns were observed at the other 2 sites. For CHW site, the OR for COVID-19 within 90 days, 90-180 days, 180-365 days and more than 365 days are 0.12 (VE =88%, 95% CI: [86-90]), 0.30 (VE =70%, 95% CI: [65-74]), 0.30 (VE =70%, 95% CI: [66-73]), and not significant (OR= 0.22, p=0.13) respectively, and the OR for MM site in the same increasing COVID-19 vaccination time windows are 0.13 (VE =87%, 95% CI: [82-91]), 0.25 (VE =75%, 95% CI: [66-81]), 0.32 (VE =68%, 95% CI: [60-74]) and not significant (OR= 0.56, p=0.47). No significance were found for subjects who received their last COVID-19 more than 365 days across all 3 sites, this may due to the small number of cases captured within study period.

The overall adjusted OR for patient received COVID-19 vaccine within 90 days was 0.14 (VE =86%, 95% CI: [84-87]), 90-180 days ago was 0.30 (VE =70%, 95% CI: [67-77]), within 180-365 days was 0.32 (VE =68%, 95% CI: [65-70]), subjects who took their last COVID-19 vaccine more than 365 days earlier was not significant (OR= 0.48, p=0.08). This further confirmed the waning effect of COVID-19 immunization published in other studies(see Figure 3).

### 3.4 Demographic Characteristics

In the study period, males had a higher risk of SARS-CoV-2 infection relative to females across all 3 study sites, CHE(OR = 1.29, 95% CI: [1.17-1.43]), CHW(OR = 1.39, 95% CI: [1.27-1.51]), MM(OR = 1.21, 95% CI: [1.02-1.44]) and overall (OR = 1.33, 95% CI: [1.26-1.41]).

Individuals with higher age are more susceptible to hospitalized COVID-19 outcome, the higher the age the higher the odds of hospitalized COVID-19 outcome. Subjects whose age is between 31-50 have higher odds of getting hospitalized COVID-19 compared to those who are under 30 with respective OR, CHE(OR = 2.86, 95% CI: [2.16-3.79]), CHW(OR = 1.89, 95% CI: [1.52-2.34]), MM(OR = 1.70, 95% CI: [1.22-2.37]) and overall(OR = 2.00, 95% CI: [1.75-2.28]). Subjects whose age is between 51-65 have higher odds of getting hospitalized COVID-19 compared to those who are under 30 with respective OR, CHE(OR = 5.29, 95% CI: [4.02-6.98]), CHW(OR = 5.43, 95% CI: [4.43, 6.67]), MM(OR = 2.57, 95% CI: [1.84-3.59]) and overall(OR = 4.31, 95% CI: [3.78-4.91]). Subjects with age greater than 65 has the highest odds of experiencing hospitalized COVID-19 outcome among our 4 age groups, the odds ratio compared to subjects whose age are under 30 are respectively 8.32 (95% CI: [6.32-10.94]), 10.45 (95% CI: [8.53-12.79]), 4.27 (95% CI: [3.08-5.92]) and 7.44 (95% CI: [6.53-8.47]) for CHE, CHE, MM and overall.

Higher Charlson comorbidity index score is associated with higher odds of hospitalized Covid outcome.(see Figure 3).

## 4 Discussion

We found that influenza vaccination was protective against hospitalized COVID-19 infection in Delta and the Omicron variant period, but with a much lower estimate of effectiveness compared with COVID-19 vaccination.

Our analysis is in agreement with previous research indicating that influenza vaccination protects against SARS-CoV-2 infection and hospitalized COVID-19[25][31][23][32][33].Our analysis differs from some studies that indicated a large degree of protection against COVID-19 from influenza vaccination. In comparison to this work, our study was conducted in a larger patient population, and we were able to control for a larger set of confounding factors, including omorbidity.

Previous studies have shown COVID-19 vaccine protection against the SARS-CoV-2 infection wanes over time[34][35][36][37][38][39][40]. This pattern is confirmed by our results.

This analysis is subject to a few limitations. Firstly, EHR data have limitations such as inaccuracy and missingness. We are likely to have missed some SARS-CoV-2 infections as hospitals across all three sites (CHE, CHW, and MM) encompass many counties and even state lines, serving many patients who might seek care or testing at other facilities. Secondly, the data was from a selected institution, which will have implications for validity outside of the study population.[41] Thirdly, although we controlled for a wide set of confounding factors, there might still be some unmeasured confounding due to higher adherence to COVID-19 mitigation measures in the vaccinated population.

## 5 Conclusion

Overall, based on our study result, influenza vaccination shown moderate effectiveness against hospitalized SARS-CoV-2 infection, but offers nowhere near the level of protection of the COVID-19 vaccines. Nonetheless, taking a flu shot is still advisable due to some extra protection and the risk of co-infection with influenza and SARS-CoV-2. Some studies have found that individuals who have COVID-19 and the flu at the same time are at much greater risk of hospitalized disease and death compared with patients who have COVID-19 alone or with other viruses[42][43][44]. Given the higher risk of hospitalized disease caused by co-infection, taking a flu shot should be encouraged to reduce the risk of COVID-related hospitalized outcomes and mortality

## Data Availability

Due to the sensitivity of real-world data, de-identified patient level are not available for public, only results, figures and tables are contained in the manuscript.

## Acknowledgments

We thank the University of Michigan Data Office for assistance with data extraction from electronic medical records. We also thank Dr. Kirsten Herold of the School of Public Health Writing Lab for her suggestions on the manuscript.

## Declarations

### Disclaimer

The funders had no role in the study design, data collection and analysis, decision to publish, or preparation of the manuscript.

